# Initial Insights from a Quality Improvement Initiative to Develop an Evidence-informed Young Adult Substance Use Program

**DOI:** 10.1101/2022.10.21.22281362

**Authors:** Jillian Halladay, Victoria Stead, Catherine McCarron, Marina Kennedy, Kyla King, Michelle Venantius, A. Carter, Sabrina Syan, Mareena Matthews, Saba Khoshroo, Myra Massey, Liah Rahman, Jacinda Burns, Kiran Punia, Emily MacKillop, Holly Raymond, James MacKillop

## Abstract

**Background:** High rates of substance misuse during emerging adulthood require developmentally appropriate clinical programs.

**Objectives:** This work outlines the development of an evidence-informed emerging adult outpatient substance use program, quality improvement process and protocol, and 1-year program insights.

**Methods:** Literature reviews, program reviews, environmental scans, and stakeholder consultations (including lived expertise) were used to develop the program. A 12-week emerging adult (17-25) measurement-based care program was developed including: 1) individual measurement-based care and motivational enhancement therapy sessions; 2) group programming focused on cognitive behavioural therapy, mindfulness, distress tolerance, and emotional regulation; 3) consults for diagnostic clarification and/or medication review; and 4) a separate education group for loved ones. A measurement system was concurrently created to collect clinical and program evaluation data at 6 time points.

**Results:** In the first year of the program, 96 young adults fully enrolled in the program (Mean age = 21 years old, 48% female gender) primarily reporting treatment targets of alcohol (70%) and cannabis (59%). Almost all patients (97%) surpassed at least one clinical threshold for co-occurring mental health disorder, with the median/mode of positive psychiatric screens being for 5 conditions.

**Conclusions:** This program demonstrates that developing an integrative evidence-informed measurement-based care young adult substance use program is feasible, though requires flexibility and ongoing monitoring to meet local needs. Patient characteristics reveal very high rates of concurrent psychiatric disorders in addition to substance use disorders.

## Introduction

Emerging adulthood, roughly operationalized as 17-25 years of age, putatively represents an “in-between” developmental period at the interface of adolescence and adulthood (1).

Relative to adolescents and adults, emerging adults experience the highest incidence and prevalence of substance use disorders (SUDs) and comorbid mental health disorders (2-4). This occurs alongside many important neurodevelopmental and psychosocial changes, including the transition from pediatric to adult healthcare systems (1, 5). Thus, emerging adults may be seeking out substance use services for the first time or transitioning to new providers with the emergence of worsening and comorbid symptoms in a system that lacks navigation support and developmentally-tailored programs (6, 7). Existing substance use services are often aimed at adults in general, and emerging adults within these programs tend to have lower motivation to change, lower engagement and retention, and, ultimately, poorer responses to treatment (6).

However, entering SUD treatment early has been related to better long-term quality of life and functioning (8), making developmentally-tailored clinical programs for emerging adults a very high priority.

To minimize the risk of emerging adults “falling through the cracks,” provincial and national governments have emphasized the need to increase integrated and concurrent treatment capabilities in all sectors of youth and adult care (9-12). However, there are gaps in the literature regarding emerging adult treatment that contributes, in part, to a lack of evidence-based emerging adult focused services (6, 7, 13). Given the unique developmental factors and sparsity of clinical research specific this age group, emerging adult substance use programs may benefit from taking a measurement-based care (MBC) approach. MBC is a contemporary approach to patient assessment, ongoing monitoring, and evaluation within healthcare systems involving routine and repeated use of standardized scales integrated into care (14). MBC has been associated with multi-faceted clinical benefits, including greater retention, engagement, and therapeutic alliances during treatment with better patient outcomes that occur more quickly and last longer than traditional care (14-19). Further, MBC data can also be used for quality improvement and research. Collectively, these benefits may help improve treatment engagement and outcomes among emerging adults accessing current substance use services while contributing to critical gaps in research to inform future improvements. However, MBC is underutilized in mental health and substance use treatment (15, 20).

The emergence and implications of substance use and comorbid mental health concerns during this critical period, compounded with the mismatch in unique developmental needs with available adult SUD programs, calls for an urgent creation of emerging adult concurrent disorders programs. This paper focuses on the development considerations of a three-year MBC emerging adult outpatient concurrent disorders pilot program for emerging adults called, the Young Adult Substance Use Program (YA-SUP). Though the target patient population for this program is “emerging adults,” consultations with emerging adults with lived expertise did not resonate with this term and preferred the term young adult (young adult used subsequently). This paper outlines the program development process, MBC system approach, program structure, initial patient characteristics in the first year of the program, and future directions.

## Methods

### Context and Eligibility

The YA-SUP is a collaboration between the Peter Boris Centre for Addictions Research (PBCAR) and the General Psychiatry Services/Concurrent Disorders Program at St. Joseph’s Healthcare Hamilton (SJHH) located in a large urban city (Hamilton, Ontario). Services are freely available to individuals 17-25 who are covered by the Ontario Health Insurance Plan (OHIP). The only additional eligibility requirement is an interest in changing substance use.

### Intervention: The Young Adult Substance Use Program (YA-SUP)

#### Program Development Process

Program development broadly followed recommendations for developing and evaluating complex interventions (21), environmental scans in health services research (22), and is consistent with the Quality Enhancement Research Initiative (QUERI) Roadmap for Implementation Quality Improvement (23). First, the YA-SUP was iteratively developed through a comprehensive environmental scan using internal and external data sources (24). Both passive (i.e., existing data, reports, guidelines, research) and active (i.e., observations, consultations) data sources were collected. Internal sources included reviewing the adult program procedures, structure, and data as well as consulting with clinicians, leadership, and administrators. A similar approach was taken with related internal clinical programs. External passive sources included reviewing current best practices for adolescent and adult substance use and concurrent disorders, local service gaps, novel research on emerging adult substance use concerns, existing adolescent and emerging adult mental health and substance use program components and structures at other institutions, and evaluated psychotherapeutic manuals. External active sources of data included stakeholder consultations with key informants including national and international experts leading research and clinical practice related to adolescent or emerging adult substance use, local emerging adults with lived expertise, and clinicians within pediatric mental health services. A foundational program model was created based on existing best practices that was presented to stakeholders updated based on feedback. See Supplementary Materials for a list of stakeholders (summary of consults available upon request).

Throughout program development, emerging adult perspectives were incorporated and amplified in several ways. First, we drew heavily on published reports and papers that have systematically collected and synthesized youth voice related to mental health care, substance use services, and transitions to the adult system. Second, we consulted with researchers and clinicians who routinely engage in co-creation and youth engagement for their services and projects. Third, in line with national standards for youth engagement (25-27), we engaged in direct youth consultations with two local emerging adults with lived experience prior to and following the initial program launch (28).

#### Environmental Scan Summary

Broadly, substance use treatment tailored to emerging adults should be grounded in harm-reduction, take a trauma-informed and concurrent disorders lens, be multidisciplinary and grounded in the biopsychosocial model, be patient-centred and driven, foster inclusivity, and focus on engagement rather than strict adherence (6, 29, 30). Emerging adults are unique, biologically, socially, and psychologically yet research looking specifically at treatments for emerging adults is sparse. However, recommendations of “meeting the patient where they are at,” being non-judgemental, and emphasizing patient self-efficacy and collaborative treatment planning are consistent with both the principles of motivational interviewing (31) and the existing practice recommendations for SUDs among adolescents and adults.

Cognitive behavioural therapy (CBT) combined with motivational enhancement therapy (MET) is the current recommended first line psychotherapeutic approach for treating both adolescent and adult SUDs (32, 33). Of note, many guidelines recommend simultaneous screening and treatment of both substance use and other mental health disorders (34) and CBT is first line psychotherapeutic treatment for common co-occurring mental health disorders like anxiety and depression in both adolescents and adults (35-41). A review of treatment for adolescent-specific substance use indicated the most promising treatments are family-based therapies, CBT, and multicomponent approaches (e.g., MET/CBT) (42). A meta-analysis of 12-20 year olds further extended support for the importance of family-based interventions, brief motivational interviewing, and MET/CBT for youth alcohol use, other drug use, and related substance use problems (43). Although they did not find effective interventions for youth cannabis use (43), evidence was limited and imprecise. Specific to motivational brief interventions, although several reviews have not found changes in frequency of use regarding cannabis use (43, 44), there are benefits for alcohol and other substance use problems among adolescents (45) and adults (46, 47) and some evidence of benefit regarding other cannabis-related outcomes among emerging adults (44). Specific to emerging adults, though limited, existing research has shown potential benefits from cognitive-behavioural and motivational interventions, integrated mental health and substance use treatment, and higher emerging adult retention when programs include CBT (6, 48, 49).

For adolescents, there is also emerging evidence for adjunctive approaches, some of which include: exercise, goal setting and progress monitoring, and third wave CBT including mindfulness, Dialectical Behaviour Therapy (DBT) skills (namely, emotion regulation), and adolescent community reinforcement approach (ACRA) (42, 50-52). ACRA combines key CBT ingredients and family-based approaches with a strong focus on substance-free reinforcements (53). ACRA has been considered part of an emerging field of behavioural economic approaches for addiction (54), which focus on minimizing demand and craving for substances, enhancing future oriented thinking and reducing impulsivity, and reducing substance-related reinforcement while maximizing substance-free reinforcements (54). Interventions for emerging adults substance use often incorporate values exploration, a behavioural economic strategy; values exercises are consistent with developmental characteristics of emerging adults who have increased self-awareness and engagement in identify exploration and formation (1, 55). Overall, these findings are in line with recommendations for emerging adults with SUDs, which primarily suggests a suite of CBT interventions, and additionally encourages incorporating social support interventions and increasing engagement in substance-free activities (6).

Although the adult healthcare system typically operates independent from the family, family-based approaches have demonstrated some of the strongest benefits for adolescent substance use (32, 43) and there appears to be continued clinical importance of family engagement during emerging adulthood (6, 56). Experts and emerging adults have also indicated that parent, caregiver, and partner involvement should be offered, while simultaneously respecting the emerging adults’ autonomy and independence, such as through separate programming (6, 29, 56, 57). There is a version of ACRA created for the loved ones called Community Reinforcement Approach and Family Training (CRAFT), which supports individuals in understanding and implementing: positive contingencies and natural consequences, positive communication and motivational strategies, and self-care strategies to improve their personal well-being (58). CRAFT is commonly recommended in family-involvement calls to action (56, 57) as CRAFT has previously been shown to increase patient engagement in treatment (59, 60), improve family functioning (58, 60), reduce the patients’ substance use (61), and improve the loved ones’ perceived empowerment (62) and mental health (63).

Lastly, it is important to ensure access to medications for both psychiatric concerns and addictions, particularly for opioid used disorder (49, 64, 65). Although it is important to simultaneously treat psychiatric disorders with medications where indicated, solely intervening with medications for non-substance related psychiatric concerns is insufficient to treat adolescent SUD (66) and other psychological and pharmacological interventions for SUDs should be delivered concurrently (67).

#### Initial YA-SUP Model

The YA-SUP’s mission is to provide young adults with the support and skills to: a) reduce the negative impact of substance use on their lives, whether that be through abstinence, reduction in use, or other harm reduction approaches; b) improve mental health and wellbeing by considering the whole-person; and c) increase engagement in substance-free activities and create a life that aligns with their values and goals. The YA-SUP values include: providing young adult centered care, creating a safe(r) space, considering the whole person, collaboration with young adults and community providers, and using evidence-based practices and contributing to evidence. The YA-SUP acknowledges the contribution of biological, psychological, and sociocultural factors in the development of addiction as well as recovery (68). This approach to treatment incorporates: a) psychological treatment including identification, psychoeducation, and treatment related to comorbid mental health concerns as well as offering psychotherapy to increase coping, identification and management of triggers, and promote ongoing recovery; b) sociocultural treatment including enhancing social support for recovery and establish a set of pleasurable substance-free activities; and c) biological and medical treatment including providing diagnostic assessments and pharmacotherapy where warranted. The program theory (21) has been simplified and visualized into a causal model and a logic model (See Supplementary Materials). The program development process has been conceptualized as a Plan-Do-Study-Act (PDSA) cycle (See Figure 1).

#### Young Adult Stream

Structurally, the YA-SUP operates around a core 12-week cycle including 2 separate streams for young adults and their loved ones. The Young Adult stream includes: 1) 5 individual sessions (the core components) grounded in MBC and MET which are supplemented by behavioural economics and ACRA interventions (e.g., values, substance-free reinforcers, quality of life and related goals); 2) near daily group programming (peripheral and adaptable components, presented as a menu of options) including CBT skills (strongly recommended as a core component), Mindfulness Based Stress Reduction, DBT informed emotion regulation and distress tolerance skills, substance related health promotion (i.e., exercise, sleep, nutrition); and 3) consults for diagnostic clarification and/or medication review and initiation.

Individual and group sessions are structured to facilitate assessment, treatment, and referrals for young adults within ∼12-week cycles (target dose: 2 sessions or groups per week, 24 sessions in 12 weeks), however, young adults in the program can still access care beyond the 12-week period. The 12-week cycle was selected to balance evidence-based research, where manualized treatments are often last 8-12 weeks, and available resources (See Figure 2). The Young Adult Stream is based on common core components of existing evidence-based manuals that were adapted prior to program launch to meet the unique needs of emerging adults. To address these multi-pronged goals, the YA-SUP has a multi-disciplinary and collaborative team which includes a Social Worker, Community Support Counsellors, Nurse Practitioner, Clinical Psychologists and trainees, Psychiatrist, and substance use and mental health researchers. Frontline staff were provided training related to all manualized sessions (e.g., CBT, MET, CRAFT). Please see Figure 3 for a summary of the YA-SUP Functional Analysis of how each program component fits into our overarching view of causal mechanisms of emerging adult substance use (based predominantly on: MacKillop (54) and Bergman, Kelly (6)).

The program implemented unique service delivery and structural characteristics recommended for emerging adult programs. Where possible, we utilize technology, such as program promotion through social media, co-creation of a program webpage, text messaging appointment reminders, and telemedicine or e-delivery of programming (6, 29, 42). Groups were delivered through telemedicine, and individual follow-ups were available via telemedicine if desired (though most chose to attend in-person). Flexible drop-in programming and short wait times, such as same-day appointments and rolling recruitment were offered (6, 13, 29). Further, in line with recommendations to include harm-reduction as a goal to treatment, clean supplies and naloxone kits are available for patients who need them.

#### Loved Ones Stream

An 8-session Loved Ones Education Group was created based on CRAFT (58, 69) and tailored to the specific needs of the loved ones of emerging adults (e.g., drawing on content from Change (70)). The sessions include: 1) Group overview, safety, and self-care; 2) understanding substance use; 3) understanding co-occurring mental health concerns; positive communication; 5) past patterns and new strategies; 6) rewards and coping with intoxications; 7) allowing negative consequences; 8) talking about treatment, review, and next steps. The specific goals for our Loved Ones Education Group are to create a sense of community, increase knowledge, and provide attendees with new strategies.

## Studying the Program

### Creating a Measurement-Based Care System

The YA-SUP was developed to be a learning health system, whereby data is collected and used to continuously update the program and respond to patient needs (23). Alongside clinical chart reviews, a priority for the YA-SUP was to embed measurement-based care (MBC) in the program. MBC refers to a structured system of assessments to promote personalized and adaptive treatment. The YA-SUP assessment (called the “battery”) evolved through literature reviews, discussion of clinically important outcomes with stakeholders, and previous batteries that have been successfully deployed in similar clinical settings (71-73).

We adopt the definition of recovery from Witkiewitz and colleagues (2020) (74), that defines recovery as *“a dynamic process of change characterized by improvements in health and social functioning, as well as increases in well-being and purpose in life*.*”* This definition supports both abstinence and harm reduction goals which have been highlighted as important aspects of emerging adult care (6, 29, 30), allowing for a person-centred approach and capturing the multidimensionality and heterogeneity of symptoms and recovery (51, 52, 67). This also follows Diagnostic Statistical Manual 5 (75) definitions of SUD remission, where the focus is on reducing problems related to use, enhancing meaningful activities, and increasing control over use rather than narrowly focusing on abstinence or reductions in substance use. It also supports expansion of clinical outcomes to include problems related to substance use such as: mental health and wellbeing; social, academic, and occupational functioning; quality of life and valued living; and engagement in substance free activities which are particularly relevant for young adults (43, 44, 55). Published reports and papers that have systematically summarized youth voices related to mental health care, substance use services, and transitions to the adult system (29, 30, 76-78) indicate improvements in quality of life is perceived as more important than substance use and mental health symptom reduction, which was echoed in stakeholder consultations. Thus, the core objectives of the YA-SUP program evaluation are to examine whether participation in the YA-SUP results in reductions in substance use and improvements in mental health and quality of life among young people with substance use concerns.

The battery was created with the aim of capturing the complexity of factors seen in emerging adults presenting to addictions services. We include questions related to (See Table 1): demographics, historical experiences, substance-use, mental health symptoms, quality of life and functioning, possible mechanisms of substance use recovery and vulnerability, and satisfaction. Emerging adults are asked to complete assessments via RedCap (79) on five occasions (with the support of a clinical research assistant): at intake, three check-in appointments approximately 4 weeks apart, and ∼12 weeks after discharge. These assessments are used to generate near real-time reports at multiple levels including personalized feedback for the young adult at each individual session, a clinician summary report to aid in conceptualizations and clinical decision making for each individual session, and a weekly program monitoring report using aggregated patient data (also supports internal data monitoring), while building a database to use for subsequent program quality improvement and research (Hamilton Integrated Research Ethics Board #12926). The clinician report includes a summary of patient demographics, frequency of substance use and readiness to change, history of trauma or head injury, and flags for whether patients surpassed clinical cut-offs for all psychiatric concerns assessed. The key components for personalized feedback reports include: 1) substance use frequency and targets for treatment; 2) quality of life; 3) SUD symptom scores; 4) anxiety and depression symptoms; and 5) personal values. Further, when iteratively adjusting the program based on clinical feedback, we follow PDSA cycle procedures by identifying key areas of improvement, creating and piloting program modifications, and evaluating the effectiveness of those changes using data from the embedded measurement system and related clinical discussions (23). See Supplementary Materials for example clinician and personalized feedback reports.

**Table 1.**
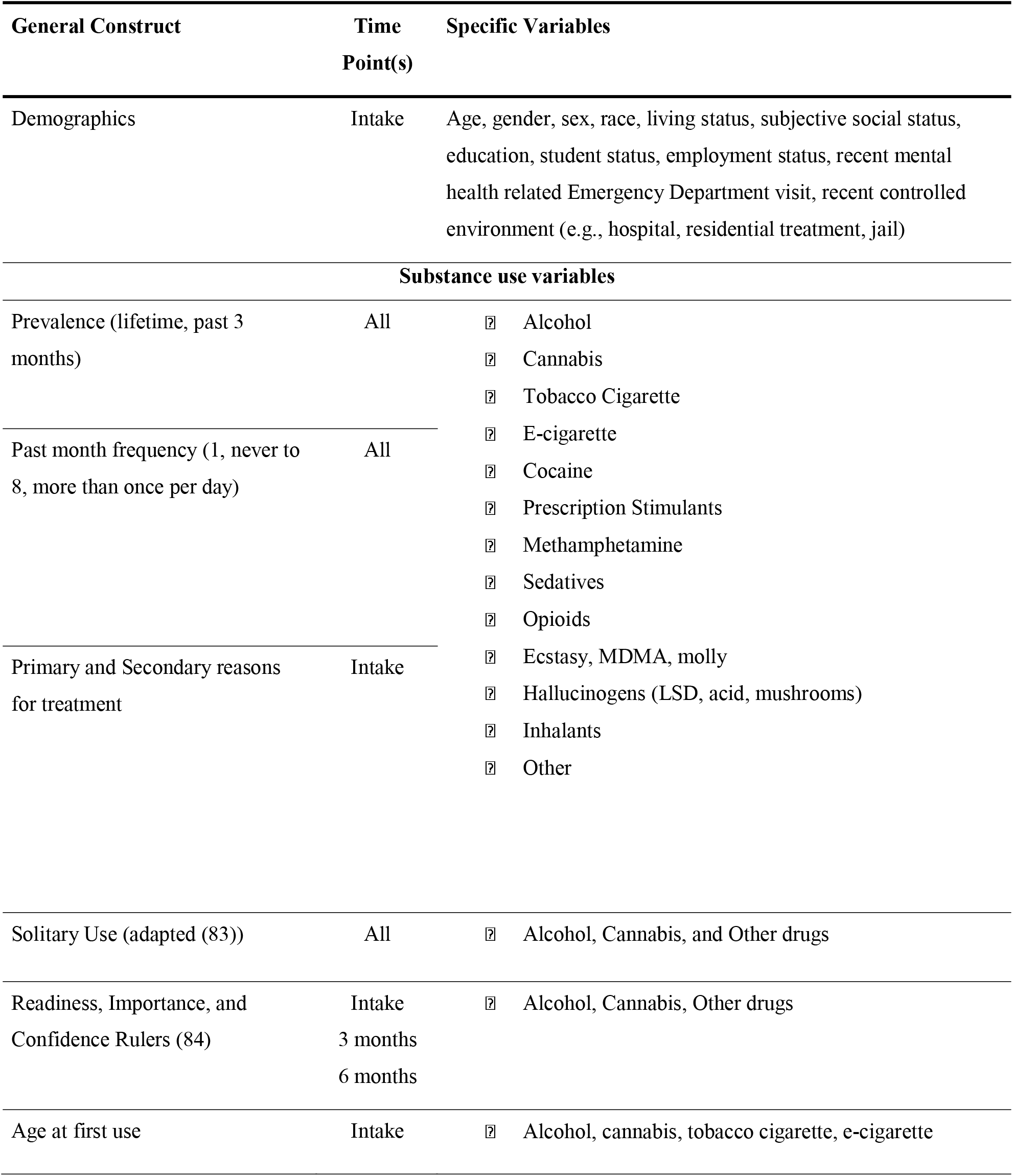
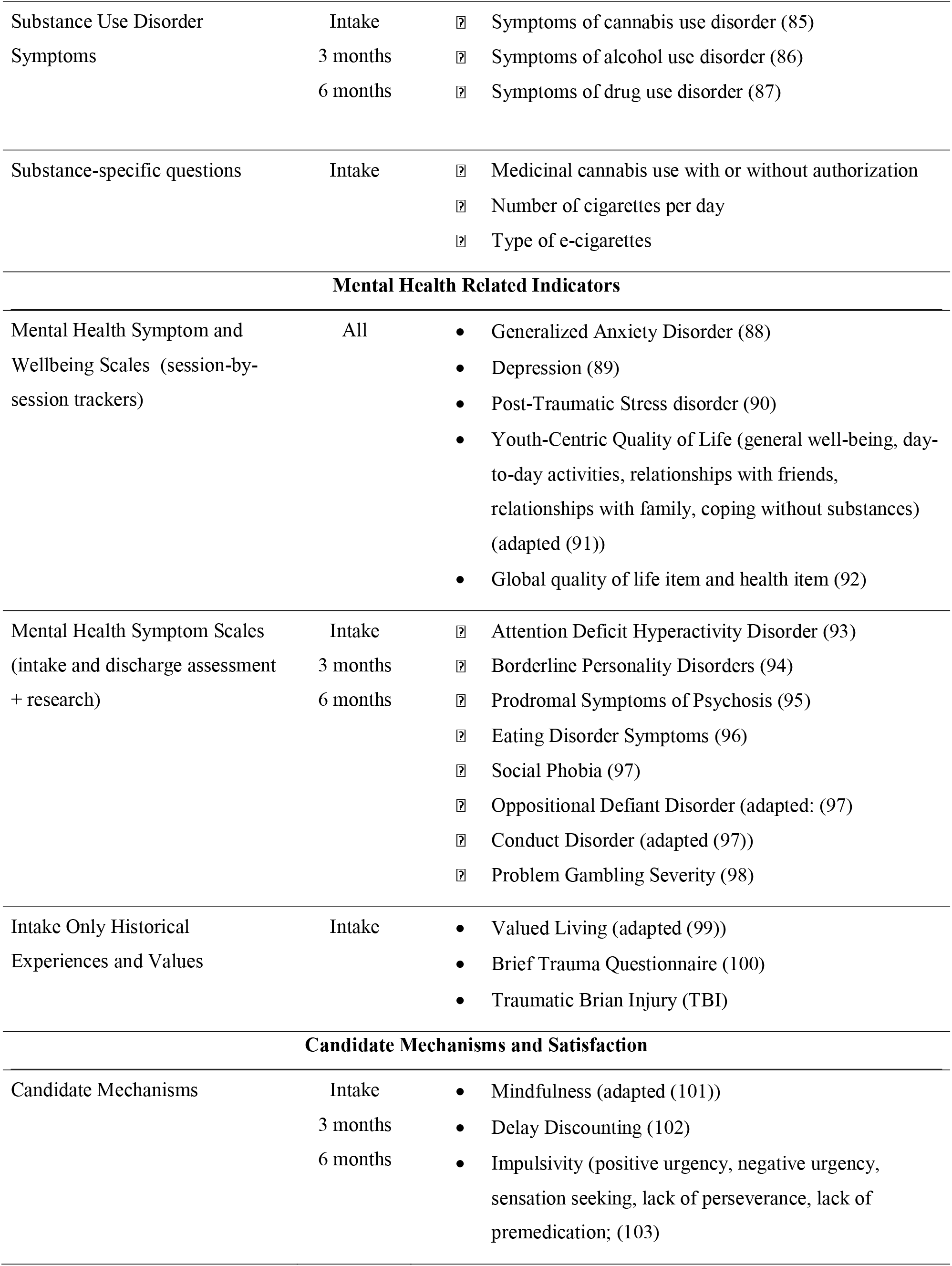
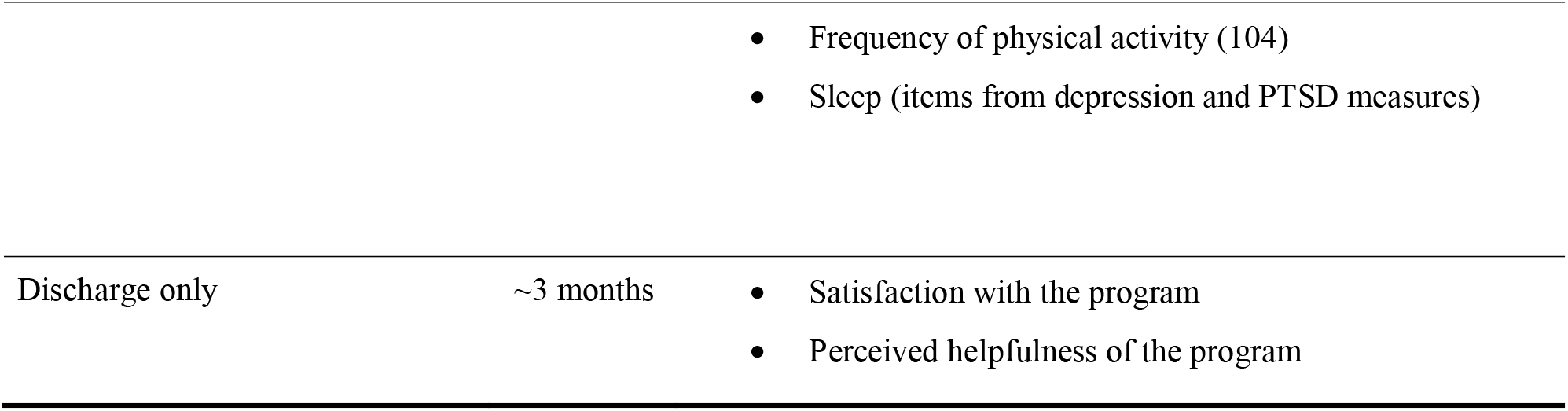
Summary of key measures

### Analysis Plan

Using an observational cohort design, the measurement system enables the program to: 1) characterize young adults accessing the program; 2) gather ongoing insight into feasibility (e.g., engagement, retention) and acceptability (e.g., satisfaction); and 3) quantify changes in substance use, mental health, and quality of life related outcomes throughout the program and at 6 months follow-up. This paper will provide detailed descriptive statistics (objective 1) regarding the clinical characteristics of patients accessing the YA-SUP in the first year, between February 22, 2021, to February 28, 2022.

## Results

During the first year of the program, 183 young adult intakes were scheduled, 124 intakes initiated, and 96 fully enrolled (i.e., completed both intake sessions) in the program (77% of intakes). For intakes, there was a 33% absentee rate and 17% did not fully enroll in program. See Table 2 for full descriptive statistics.

**Table 2.** Demographic and Clinical Patient Characteristics (n=96)

| *Socio-demographics* | **Mean (SD) or % (n)** |
| --- | --- |
| Age | 21.1 (2.6); range 16-25 |
| White | 83.3% (80) |
| Female | 54.2% (52) |
| Gender |  |
| Woman | 45.8% (44) |
| Man | 45.8% (44) |
| Non-binary, don't know, prefer not to answer | 8.2% (8) |
| Living Situation |  |
| Living with family | 54.2% (52) |
| Living independently | 16.7% (16) |
| Living with a partner, roommate(s), or in residence | 26.0% (25) |
| Living in a group home, shelter, or homeless | 3.1% (3) |
| Subjective Socioeconomic Status |  |
| Not enough to pay bills | 11.5% (11) |
| Enough to pay bill, but have to cut-back | 26.0% (25) |
| Enough to pay bills without cutting back | 25.0% (24) |
| Enough for extras | 37.5% (36) |
| Student |  |
| Not a student | 66.7% (64) |
| Part time or less | 11.4% (11) |
| Full time | 21.9% (21) |
| Employment |  |
| Not employed | 42.7% (41) |
| Casual or part time | 34.4% (33) |
| Full time | 22.9% (22) |

| *Substance Use* | **Any use in the past 3 months**<br>% (n) | **Frequency of past month use<sup>a</sup> mean (SD)** |
| --- | --- | --- |
| Alcohol | 84.4% (81) | 4.3 (2.1) |
| Cannabis | 87.5% (84) | 6.5 (2.4) |
| Tobacco Cigarettes | 75.0% (72) | 6.0 (2.6) |
| E-cigarettes | 58.3% (56) | 6.2 (2.5) |
| Cocaine | 31.3% (30) | 3.2 (2.1) |
| Prescription stimulants | 11.5% (11) | 3.3 (2.6) |
| Methamphetamine | 7.3% (7) | 2.7 (2.5) |
| Sedatives or Tranquilizers | 13.5% (13) | 2.5 (1.1) |
| Opioids | 13.5% (13) | 2.6 (2.0) |
| Ecstasy, MDMA, molly | 8.3% (8) | 2.0 (0.8) |
| LSD, Acid, or mushrooms | 18.8% (18) | 2.3 (1.1) |

| *Readiness Rulers<sup>b</sup>* | **Readiness**<br>**Mean (SD)** | **Importance**<br>**Mean (SD)** | **Confidence**<br>**Mean (SD)** |
| --- | --- | --- | --- |
| Alcohol | 6.5 (3.3) | 6.3 (3.5) | 6.1 (3.3) |
| Cannabis | 4.8 (3.7) | 5.1 (3.8) | 4.3 (3.5) |
| Other Drugs | 7.5 (3.2) | 6.9 (3.8) | 7.0 (3.2) |

| *Mental Health Indicators* | **Mean (SD)** | **Clinical**<br>**Threshold** | **% (n) surpassing threshold** |
| --- | --- | --- | --- |
| Quality of Life (0-50) | 20.1 (10.7) | N/A | N/A |
| Trauma Exposure (0-11) | 3.3 (2.4) | 1+ | 90.6% (87) |
| Traumatic Brain Injury (0-5) | 1.4 (1.4) | 1+ | 65.6% (63) |
| Alcohol Use Disorder (0-40)<sup>b,c</sup> | 17.5 (10.1) | Moderate: 8-14<br>High: 15+ | Moderate: 20.8% (20)<br>High: 47.9% (46) |
| Cannabis Use Disorder (0-32)<sup>b</sup> | 18.2 (8.5) | Moderate: 8-11<br>High: 12+ | Moderate: 7.3% (7)<br>High: 66.7% (64) |
| Drug Use Disorder (0-44)<sup>b</sup> | 19.2 (10.9) | High: 15+ | 18.8% (18) |
| Generalized Anxiety Disorder (0-21) | 14.9 (5.4) | Moderate: 10-14<br>Severe: 15+ | Moderate: 20.8% (20)<br>Severe: 60.4% (58) |
| Depression (0-27) | 18.0 (6.2) | Moderate: 10-14 | Moderate: 13.5% (13) |
|  |  | Mod-Severe: 15-19 | Mod-Severe: 31.3% (30) |
|  |  | Severe: 20+ | Severe: 44.8% (43) |
| Social Anxiety Disorder (0-10) | 6.4 (2.9) | 10+ | 18.8% (18) |
| PTSD (0-80) | 44.2 (20.3) | 42+ | 54.2% (52) |
| Psychosis (0-16) | 7.3 (4.2) | 6+ | 66.7% (64) |
| Borderline Personality Disorder (0-10) | 6.9 (2.6) | 7+ | 61.5% (59) |
| ADHD (0-24) | 15.3 (5.4) | 4+ of select variables | 61.5% (59) |
| Oppositional Defiant Behaviours (0-12) | 5.0 (3.2) | 7+ | 36.5% (35) |
| Conduct Behaviours (0-22) | 3.5 (3.6) | 7+ | 15.6% (15) |
| Surpassing any non-SUD mental health threshold | 4.8 (2.3) | 1+ | 96.9% (93) |

<sup>a</sup>Coded: 1=not in the past 4 weeks, 2=1 day in the past 4 weeks, 3=2 to 3 days in the past 4 weeks, 4=1 to 2 days per week, 5=3 to 4 days per week, 6=5 to 6 days per week, 7=once per day, 8=more than once per day.
<sup>b</sup>Among those endorsing any use in the 3 months prior to intake.
<sup>c</sup>Due to an administrative error, response options were slightly different from the original measure. See SMI for details.

Of young adults who fully enrolled in the first year (n=96), patients were on average 21 years of age and 46% were women. Most young adults were coming to the program for their alcohol (67%), cannabis (60%), cigarette (37%), and/or cocaine use (27%) (Supplementary Materials). There were high levels of comorbid types of SUDs (Figure 4A). Almost all young adults (97%) surpassed at least one clinical threshold for co-occurring non-substance mental health symptoms; on average, young adults surpassed 5 different co-occurring clinical thresholds (Figure 4B). In terms of specific conditions, 76% endorsed moderate to severe depressive symptoms, 81% moderate to severe generalized anxiety symptoms, and there was near universal trauma exposure (91%) with 54% surpassing PTSD thresholds (Figure 4C). Notably, 46% surpassed cut-offs for GAD, Depression, BPD, and PTSD. On average, young adults were reporting 4 out of 10 across various youth-specific domains of quality of life at intake (Supplementary Materials). Further, 54% of the patients were living with their family and 26% with others (partners, friends, roommates), highlighting the importance of the Loved Ones Education group.

The program also delivered three cycles of the Loved Ones Education Group, with all attendees identifying as parents or caregivers of young adults with substance use concerns. Of the attendees (n=∼33), a majority of their young adult loved ones were not in treatment (76%) and predominantly using cannabis (82%), alcohol (48%), and/or cigarettes (24%).

## Discussion

The Young Adult Substance Use Program (YA-SUP) represents the combination of best practices, contemporary research, and stakeholder input, while operating within the parameters of available program resources. This program demonstrates that developing an integrative evidence-informed measurement-based care young adult substance use program is feasible, though requires flexibility and ongoing adaptations to meet local needs. In the first year of the program, alcohol and cannabis use were the most common reasons for treatment and emerging adults presented with a high degree of comorbidity, particularly regarding internalizing mental health disorders and near-universal trauma exposure. The level of complexity and comorbidity among young adults presenting to the program was higher than anticipated. By taking a measurement-based care and learning health system approach, we have been able to simultaneously monitor who our program is serving and how our program is doing to inform immediate improvements to the program and contribute to gaps in research.

We made a number of notable, iterative, systematic modifications to the delivery and evaluation of the program during the first year (23). These adaptations included: separating content initially developed for single sessions into multiple sessions (e.g., intake, CBT identifying and challenging thoughts); adjusting group structure (e.g., reducing number of young adult group options) and content (e.g., adding more co-occurring mental health content for loved ones); clarifying, adding, and streamlining questions in the assessment battery and refining clinician reports and patient personalized feedback based on these assessments (i.e., the measurement system); and expanding the clinical team (notably, hiring a community support worker focused on community outreach, engagement, collaboration, and capacity building). By reviewing young adult characteristics from the battery alongside implementation outcomes (such as reach, uptake, and dose), clinical insights from the care team, and follow-ups with young adults, we are continuing to iteratively improve the program. Specifically, like other similar programs, we have seen relatively high levels of service disengagement early in the program (80-82). By leveraging our measurement system, we have been able to explore predictors of retention and engagement in our population. Accordingly, the clinicians are leading the development and implementation of pilot programming focused on individualized harm reduction, trauma, and group readiness (including related anxiety, severe emotion dysregulation, and behavioural activation and planning).

Generalizability regarding specific program components and patients is limited due to the program being created for and implemented in a single city. Further, several best practices and stakeholder suggestions have not yet been incorporated, in part due to resource limitations. Thus, future considerations for our programs and others include: expanding flexibility in the referral pathways, street outreach, timing, and location of services; expanding program offerings to focus on other areas of health and functioning (e.g., occupational support, housing, exercise, finance, substance-free activities); increasing access to longer term psychopharmacotherapy monitoring; mutual support or weekly structured relapse prevention groups; and community capacity building for adolescent services in collaboration with other local youth substance use services.

Overall, this paper summarizes: 1) the rationale and key considerations for the development of an outpatient program for emerging adult co-occurring substance use and mental health concerns; 2) the development and implementation of an embedded measurement system that provides the structure for future program evaluation; and 3) initial patient characteristics, clinical insights, and iterative clinically-reactive adaptations following year 1 of program launch. This program overview can help inform the development and evaluation of future tailored programs.

## Data Availability

All data produced in the present study are available upon reasonable request to the authors

## Acknowledgements

We thank the Boris Family for their generous donation that made this program possible. We also want to thank all of the research, clinical, and young adult experts who consulted on program development including: Brandon Bergman, PhD; Khrista Boylan, MD, FRCP(C), PhD; Penny Burley, CRPO; Kim Corace, PhD; Andrew Costa, PhD; Sarah Feldstein Ewing, PhD; Melissa Griffin, PhD; Taylor Hatchard, PhD; Lisa Hawke, PhD; Joanna Henderson, PhD; Ellen Lipman, MD, FRCP(C), MSc; Lisa Jeffs, MA; Madeleine Luvisa, BSW; Leslie Martin, MD; Mackenzie Mawson, BScN, RN; Catherine McCarron, RSW, MSW; Bob Miranda, PhD MEd; Catharine Munn, MD, FRCP(C), MSc; Jim Murphy, PhD; Dawn Pierce, RN; Tim O’Shea, MD; Christine Squires, Community Support Worker; Elizabeth Osuch, MD; John Westland, MSW. We would also like to thank all the clinical and research trainees and staff who have worked with the YA-SUP. Lastly, we would like to thank the patients and families who trust us with their care.

