## Supplementary Materials for "Initial Insights from a Quality Improvement Initiative to Develop an Evidence-informed Young Adult Substance Use Program"

**Supplementary Materials A-H**

A. List of Researcher and Clinician Stakeholder Consultations

B. Program Causal Model

C. Program Logic Model

D. Detailed rationale of young adult program ingredients

E. Details overview of Loved Ones Education Group

F. Example Clinician Summary

G. Example Personalized Feedback Report

H. Additional Young Adult Characteristics

I. AUDIT items

**Supplementary Materials A**

**A. List of Researcher and Clinician Stakeholder Consultations**

Consultations (including research and clinical consultations) were done via Zoom August 2020 - November 2020 and typically included:

- An overview of the most up to date program goals and structure (iteratively developed with accumulating literature and consultations)
- Feedback on the current goals and structure
- Suggestions for improvements or adaptations
- Suggestions for specific programming or program components/organization

**Research Consultants:**

- **Brandon Bergman, PhD** Assistant Professor at Harvard Medical School and Associate Director at the Recovery Research Institute
- **Andrew Costa, PhD** Schlegel Research Chair in Clinical Epidemiology & Aging, Department of Health Research Methods, Evidence, and Impact at McMaster University
- **Sarah Feldstein Ewing, PhD** Department of Psychiatry, division of Clinical psychology in the School of Medicine at Oregon Health and Science University
- **Lisa Hawke, PhD** Project Scientist in the Margaret and Wallace McCain Centre for Child, Youth & Family Mental Health and a Cundill Scholar with the Cundill Centre for Child and Youth Depression at CAMH. She is also an Assistant Professor at the University of Toronto Department of Psychiatry and an Associate Member of the Yeates School of Graduate Studies at Ryerson University.
- **Joanna Henderson, PhD** Director of the Margaret and Wallace McCain Centre for Child, Youth & Family Mental Health and Interim Implementation Director of the Cundill Centre for Child and Youth Depression at CAMH. She is also Senior Scientist in the Child, Youth and Emerging Adult Program at CAMH, and Associate Professor in the Department of Psychiatry at the University of Toronto
- **Jim Murphy, PhD** Department of Psychology at the University of Memphis

**Clinical and Young Adult Advocate and Experience Consultants:**

- **Khrista Boylan, MD, FRCP(C), PhD** Clinical Researcher (Child Psychiatrist) in Department of Psychiatry and Behavioural Neurosciences and Clinical Epidemiology and Biostatistics at McMaster University
- **Penny Burley**, **CRPO** Executive Director at Alternatives for Youth Hamilton in Hamilton Ontario
- **Kim Corace, PhD** Clinical Psychologist and Director of Program Development and Research within theSubstance use and Concurrent Disorders program at The Royal Ottawa Mental Health Centre and Professor in the Department of Psychiatry within the Faculty of Medicine at the University of Ottawa
- **Melissa Griffin, PhD** Advanced Practice Clinical Leader for the Youth Addiction and Concurrent Disorders Service (YACDS), The Substance Abuse Program for African Canadian and Caribbean Youth (SAPACCY), Child, Youth and Family Service, and Youth Urgent Care Clinic at the Centre for Addiction and Mental Health in Toronto, Ontario
- **Taylor Hatchard, PhD** Psychologist at the Youth Wellness Centre in Hamilton Ontario and Assistant Professor within the Department of Psychiatry and Behavioural Neurosciences at McMaster University
- **Lisa Jeffs, MA** Program manager of the Youth Wellness Centre in Hamilton Ontario
- **Ellen Lipman, MD, FRCP(C), MSc** Director of the Offord Centre of Child studies, Head of the academic Division of Child Psychiatry, and Chief of the Department of Child Psychiatry at McMaster Children’s Hospital and McMaster University
- **Mackenzie Luvisa, BSW** Youth Wellness Centre in Hamilton Ontario
- **Mackenzie Mawson, BScN, RN** Youth Wellness Centre in Hamilton Ontario, Hamilton Health Sciences
- **Catherine McCarron**, RSW MSW Manager of the RAAM (Rapid Access Addiction Medicine) Clinic, The Capacity Building Team & Community Psychiatry Clinic - Bridge to Recovery, Concurrent Disorders Outpatient, Borderline Personality Disorder Services, Rapid Consultation & General Psychiatry at St. Joseph’s Healthcare Hamilton
- **Leslie Martin**, **MD** General Internist, Assistant Professor, Department of Medicine, Division of General Internal Medicine at McMaster University
- **Bob Miranda, PhD MEd** Director of the Vista Clinic at Bradley Hospital and Professor of Psychiatry and Human Behavior, Professor of Behavioral and Social Sciences within the Center for Alcohol and Addiction Studies at Brown University
- **Catharine Munn, MD, FRCP(C), MSc** Assistant Dean of Resident Affairs, Postgraduate Medical Education (PGME), Lead of Professor Hippo-on-Campus Student Mental Health Education Program, and Associate Professor within Department of Psychiatry and Behavioural Neurosciences at McMaster University. Clinical experience in University Student Wellness Centre Psychiatry.
- **Tim O’Shea, MD** Internal Medicine Specialist, Member of the Shelter Health Network in Hamilton, Ontario, Associate Professor, Department of Medicine, Division of Infectious Diseases at McMaster University
- **Elizabeth Osuch, MD** Rea Chair of Affective Disorders, founder of the first Episode Mood and Anxiety Program (FEMAP), cross-appointed In the Department of Medical Biophysics and is part of the Lawson Research Health Institute in Neuroimaging in Mental Health at Western University
- **Dawn Pierce**, **RN** Registered Nurse in the Concurrent Disorders Outpatient program at St. Joseph’s Healthcare Hamilton
- **John Westland, MSW** Social Worker in the Adolescent Substance Abuse program at SickKids Hospital in Toronto, Ontario

**B. Causal Model (**informed by Ross, Vingilis (1))

**Young adults with substance use problems with and without mental health problems.**

Young adults self-**refer** or get referred to the YA-SUP.

Young adults engage in a **comprehensive assessment and brief intervention** during the first two intake sessions.

**PHASE 1: Rapid Access and Assessment**

**PHASE 2: Connections and Treatments to Reduce Symptoms and Improve Functioning**

Young adults reduce their harm from substances, experience **improvements in functioning and wellbeing**, and increase engagement in substance free activities and supports. Young adults continue to engage in recovery-oriented supports.

Young adults are **triaged** and directed towards needed services (internal and external referrals).

Young adults engage in **treatment.** The core treatment is offered in 12-week cycles grounded in the **YA-SUP functional analysis** of young adult substance use.

**C. Logic Model**

| **NEEDS** | **INPUTS** | **ACTIVITIES** | **OUTPUTS** | **OUTCOMES** |
| --- | --- | --- | --- | --- |
| **CLIENTS**:   - Evidence-based concurrent disorders care:   - Biopsychosocial Assessment   - Measurement Based Care (MBC) and tailored treatment planning   - Interventions – individual and group   - Consultation: diagnostic clarification and medication initiation   - Crisis care management - Meaningful activities - System navigation   **TEAM**:   - Clarity of team processes - Optimal client flow - Communication of processes (e.g., manualized procedures and training, MBC reports) - Multidisciplinary collaboration - Support for crisis management - Opportunities for professional development   **SYSTEM**:   - Clear expectations about YASUP services - Clear referral pathways (for community, providers, and centralized intake staff) - Follow-up consultation and longer-term services (e.g., shared care) - Decreased use and burden on emergency services | **TEAM**:   - Clinical Director and Manager Support - Research support - Admin support - Psychiatry - Social Worker - Community Support Counsellors - Nurse Practitioner - Clinical Psychology - Research Staff - Central Intake (CONNECT) - Hospital - Electronic Medical Record - IT and HR   **SYSTEM**   - SJHH and PBCAR - Adult Concurrent Disorders Outpatient Team - Hospital Concurrent Disorders Capacity Building Team   **INFRASTRUCTURE**   - Office Space: SJHH within CD outpatient (clinical) and PBCAR (research) - Computer, telephone, internet - Office supplies - Clinical supplies - MBC standardized system (RedCap, Battery) - Virtual platform | **PROGRAM DEVELOPMENT:** Conducting literature review, consultations with stakeholders, hiring, onboarding and training of clinical staff, creating intake process, developing a measurement system, assessing and identifying level of substance use and mental health symptomatology, and ongoing review of program.  **PATIENT INTAKE**:   - Eligibility – young adults (17-25) considering making changes to their substance use - CONNECT Screening (centralized intake) or internal screening for internal direct referrals - Intake Part A, rapid access with opportunities for same day appointments (biopsychosocial assessment) with MBC battery - Possible triage/referral to other services to direct youth to needed services (internal & external) - Intake Part B, 1 week later (‘fully enrolled’; MBC & MET)   **ASSESSMENTS:**   - Biopsychosocial intake assessment - Repeated standardized electronic MBC assessments (intake, 1 month, 2 months, 3 months, 6 months) - Consults: Psychiatry, Nurse Practitioner, Neuropsychology   **YOUNG ADULT STREAM TREATMENT:**   - Four, structured individual MBC and motivational enhancement therapy (MET) sessions (1 week and 1, 2, 3 months). Opportunity for additional individual sessions contingent on group participation. - Virtual Groups (almost all drop-in): CBT skills, DBT skills, mindfulness, health promotion   **EDUCATION FOR:**   - Patients and their loved ones - Care providers and researchers - Community partners   **EVALUTATIONS**   - Research projects - Program monitoring and evaluation   **ADVOCACY AND COLLABORATION**:   - Co-developing grants with youth to embed more integrated youth engagement in research and program development - Community outreach to enhance shared, collaborative care | **CLIENTS**:   - # Intakes - # Check-ins - # Groups - # Consults - # Loved Ones   **TEAM**:   - # Education sessions - Weekly clinical and patient flow team meetings - Bi-weekly research meetings   **COMMUNITY**:   - # Community presentations, meetings, and town halls   **SYSTEM:**   - # Externals service usage (e.g., emergency, inpatient, other SJHH services) | **SHORT TERM:**   - Publish program development and measurement system plan, alongside initial patient descriptive data - Increased access to care: short wait times to first contact(s) - Increased commitment to treatment and motivation to change among emerging adults with substance use concerns - Improvements in quality of life - Increased support to families and loved ones - High rates of satisfaction from participants   **MEDIUM TERM:**   - Reduced substance use and psychiatric symptoms - Increased engagement in substance free alternative activities including exercise, mindfulness, school/education, work/employment, and other leisure - Adapt programming based on participant feedback and engagement to improve programming   **LONG TERM:**   - Reduced rates of emerging adults “falling through the cracks” - Reduced need for acute care (i.e., emergency visits and hospitalizations for substance use or mental health concerns) - Reduced chronic illness due to substance use - Increases in stable housing and employment - Building and disseminating a model of care for emerging adult substance use to replicate and/or improve and build upon for future programs |

**D. Detailed rationale of young adult program ingredients**

1. **Comprehensive biopsychosocial assessment**

In line with summary clinical texts (2), the intake assessment for the YA-SUP is informed by the biopsychosocial model of addiction (3). It includes a comprehensive multidimensional assessment of substance use, physical health, mental health, socio-demographic risk and protective factors (e.g., family history, exposure to trauma, sex and gender, race and ethnicity), psychosocial risk and protective factors (e.g., educational and occupational functioning, family functioning, peer relations, justice involvement, recreation), and treatment history. Through the course of the assessment, a clinical formulation will provide insight into the presenting problem, predisposing factors, precipitating factors, perpetuating factors, and protective factors (4). Based on this assessment, emerging adults may be referred to an YA-SUP Nurse Practitioner and/or Psychiatrist to receive a diagnostic assessment and/or pharmacotherapy for mental health and/or substance use concerns. They may also be referred to a Clinical Psychologist for a neuropsychiatric evaluation. This intake assessment was predominantly informed by existing intake procedures in the Adult Concurrent Disorders Outpatient Program, at the Youth Wellness Centre, and the extended psychosocial assessment of young people (e.g., headspace version of HEADSS) (5).

2. **Brief Motivational Intervention & Substance Free Activity Session**

Brief motivational interventions (BIs) for adolescents and emerging adults have shown positive effects for alcohol related outcomes and general substance use related problems (6-8). Effects of BIs for cannabis use outcomes are varied with no consistent benefits for frequency (6) but some evidence of benefit on increased abstinence and reductions in symptoms of cannabis use disorder among emerging adults (9). Using behavioural economic approaches for addiction, there is emerging evidence of additional benefits of supplementing BIs with substance free activity sessions (SFAS) such as values based exercises (10, 11). Additionally, given poor treatment engagement among emerging adults (12, 13), we want to ensure all emerging adults who complete a full intake (intake assessment followed by BI 1 week later) will receive evidence-based treatment. By utilizing motivational and developmentally tailored approaches, the brief intervention during the intake predominantly aims to increase readiness to change and commitment to treatment.

The BI in the YA-SUP intake session includes all the main BI ingredients identified in a systematic review of cannabis BIs for emerging adults, including: (1) *motivational interviewing*; (2) *pros and cons*; (3) *personalized feedback* including peer normative feedback; (4) *values* and life goals; and (5) *goal setting*. Specifically, the BI was predominantly adapted from previous piloted and evaluated manuals an UpToDate Recommendations (14). One of the manuals used was a BI for university student alcohol and cannabis use, which incorporated MET/MI approaches, a values activity, and a discussion co-occurring mental health concerns (15). This manual was updated and supplemented with a substance-free-activity session (SFAS) informed by BIs with SFAS for adults and emerging adults (10, 11). The set of SFAS components incorporated include: personalized feedback and discussion from the *Valued Living Questionnaire*, discussion of *substance-free recreational activities*, and *goal setting* related to values and non-substance free activities in addition to substance specific goals. The SFAS and goal setting was further informed by the **happiness scale and goals of counseling session** from ACRA (16) but instead of using the ACRA Happiness Scale (16 items) we use the MyLifeTracker (5 items) (17).

3. **Individual MET sessions**

MET combined with CBT is the current recommended first line psychotherapeutic approach for treating both adolescent and adult substance use disorders (18, 19). The BI-SFAS embedded into the intake session will serve as the first of four motivational enhancement therapy (MET) sessions. Follow-up MET sessions are done at ~4, 8, and 12 weeks post-intake. There is some evidence suggesting spreading out one-on-one MET session can improve treatment engagement and retention among youth (20). The MET sessions were adapted from the following evaluated manuals: 1) a 2-session MET interventions for adolescent substance use (21); 2) a 4-session MET intervention used among youth with AUD during Project MATCH (MATCH); and 3) an in house contemporary 4-session MET intervention for individuals with Alcohol Use Disorder (22).

4. **Group Psychotherapeutic Approaches**

In general, group psychotherapeutic approaches for individuals with substance use with and without comorbid psychiatric concerns have shown to increase likelihood of abstinence in the short and longer term and improvements in psychological health, particularly when compared to no intervention (23, 24). In a recent meta-analysis, most outcomes were not significantly different when comparing group to individual or other active treatments (24). This evidence in combination with reduced costs for group interventions, provides support for delivering group psychotherapy for the YA-SUP. Groups can also provide an opportunity to provide a safe setting for learning and refining skills – especially social skills which are often central to CBT and substance use treatment. This is particularly important for emerging adults, as cognitive skills during this developmental period are influenced by socioemotional contexts (25). Further, YA-SUP manuals drew more heavily on existing adolescent manuals (compared to adult) given the continued development of the prefrontal cortex into mid-20s, potential negative impact of substance use on neuromaturation (26), and insight from local youth programs of better emerging adult outcomes when switching from adult to adolescent manuals.

| **Young Adult Group Psychotherapy (implemented)** | | |
| --- | --- | --- |
| Group | Session | Mechanism |
| Mind-Drug Connection (CBT core, sequential sign-up) | Understanding your substance use | Functional analysis |
|  | Identifying problematic thoughts | Cognitive triggers |
|  | Challenging problematic thoughts | Cognitive Restructuring |
|  | Coping with urges and learning refusal skills | Coping with urges |
|  | Planning for difficult situations | Relapse prevention |
| Mind-Drug Connection (CBT- drop-in) | Taking control of your anger | Interpersonal skills |
|  | Problem solving | Coping with daily life and decision making |
|  | Being assertive | Interpersonal skills |
|  | Communication skills | Social facilitation |
|  | Exploring and creating your social support network | Social facilitation |
|  | Identifying and scheduling pleasant activities | Substance free reinforcement (CBT & Behavioural Economics) |
|  | Living your values | Substance free reinforcement (CBT & Behavioural Economics) |
| Balancing Emotion and Mind (DBT skills– drop-in) | Why and how to do the opposite of how you feel (Opposite Action) | Positive Coping: Emotion Regulation |
|  | Maintaining emotional control (PLEASE skill) |  |
|  | Stopping an impulsive behaviour in its tracts (STOP skill) | Positive Coping: Distress tolerance |
|  | Thinking your way out of an impulsive behaviour (Pros and Cons Skill) |  |
|  | Turning your attention away from a negative emotion until you can resolve it (ACCEPTS skill) |  |
|  | Reducing the intensity of negative emotions by using your senses (Self Soothe skill) |  |
|  | Quickly reducing the intensity of negative emotions by changing your body physiology (TIPP skill) |  |
| Mindfulness (drop-in) | 6 session Mindfulness-Based Stress Reduction adapted program BREATH: Body, Reflections, Emotions, Attention, Tenderness, and Habits | Positive Coping: Mindfulness |
| Levelling up your health (drop-in) | Exercise, nutrition, sleep | Physical health, cognition, and emotion regulation |

**CORE: Cognitive Behavioural Therapy Group.**

CBT (alongside MET) is the current recommended first line psychotherapeutic approach for treating both adolescent and adult substance use disorders (18, 19), common co-occurring psychiatric disorders (27-31), and various combinations of CBT-related approaches have been recommended for emerging adults specifically (13). CBT not a single intervention but a family of interventions. Of note, mindfulness, ACRA, ACT, and DBT are considered third wave CBT and thus there are many shared modules and overlapping themes across various CBT approaches. Overall, the main targets of CBT for substance use include enhancing: coping, problem solving, decision making, interpersonal relationships, substance free activities, and understanding and control over one’s internal and external environment (19).

In the process of selecting the YA-SUP CBT modules, the common ingredients across various published and evaluated protocols alongside local manuals for substance use and concurrent disorders for adolescents and adults were identified. Manuals compared included:

- Motivational Enhancement Therapy and Cognitive Behavioral Therapy for Adolescent Cannabis Users. Cannabis Youth Treatment (CYT) Series (SAMHSA) (21, 32)
- C-SMART Concurrent disorders (Depression & CUD) for emerging adults (CAMH) (33)
- The adolescent community reinforcement approach for adolescent cannabis users (SAMHSA) (16)
- Cognitive-behavioral coping skills therapy manual: A clinical research guide for therapists treating individuals with alcohol abuse and dependence (34) and related/updated Coping Skills Training (35)
- Structured Relapse Prevention Concurrent Disorders (CAMH) (36)
- Crossing Paths: Cognitive Behavioural Therapy for Depression, Anxiety & Substance Use Problems (ATRC) (37)
- Project Combine: Combined Behavioral Intervention (38)
- Mind vs Mood: Changing our thoughts & behaviours using CBT strategies (YWC) (39)
- Network support for alcohol treatment: Mechanisms and effectiveness (40)

| **Core Topics from review of manuals** | |
| --- | --- |
| **1** | **Functional analysis** **and intro to CBT triangle** |
| **2** | **Identifying thoughts with a focus on substance use** |
| **3** | **Managing thoughts of substance use and negative thinking** |
| **4** | **Coping** **with cravings and urges** |
| **5** | **Problem solving** |
| **6** | **Refusal Skills** |
| **7** | **Assertiveness Training** |
| **8** | **Anger awareness and management** |
| **9** | **Communication Strategies** |
| **10** | **Social Support Networks** |
| **11** | **Pleasant activities and scheduling** |
| **12** | **Planning for emergencies and lapses** |
| ***** | **Weekly Planning Drop-in Groups** |

**Adjunctive Group Programming**

**Mindfulness Based interventions** have shown benefits for adult substance use outcomes (mainly mindfulness-based relapse prevention) (41, 42), emerging adult mental health outcomes (43, 44), and has shown preliminary evidence of effectiveness for adolescent substance use (45). Thus, the YA-SUP includes a series of 6-mindfulness modules based on a manualized and tested youth mindfulness program called Learning 2 Breath (46). Typical mindfulness interventions are often 12 weeks in duration, however, the meta-analysis of mental health outcomes among emerging adults showed no significant subgroup differences between shorter or longer interventions (44).

**DBT Emotion Regulation and Distress Tolerance Skills Sessions.** Dialectical Behaviour Therapy (DBT) is a third-wave CBT intervention the includes 4 modules: mindfulness, distress tolerance, emotional regulation, and interpersonal effectiveness. Although limited, there is some evidence of the benefits of DBT skills for substance use disorders among adults (47, 48) and many stakeholders suggested and advocated for the incorporation of DBT into the program. As noted above, many CBT and CBT related interventions share commonalty. Thus, to minimize overlap and maximize resources, we:

- Omitted DBT interpersonal effectiveness modules and use CBT modules related to communication, assertiveness, and refusal skills;
- Omitted mindfulness DBT skills sessions and use Learning 2 Breath as the manualized mindfulness intervention;
- Include emotion regulation DBT skills sessions (e.g., opposite action, PLEASE). Of note, 1) ‘check the facts’ will not be included as CBT modules include 2 sessions on identifying and challenging thoughts; 3) ‘Paying attention to positive events’ is incorporated into all groups by incorporating a moment of reflection on positive events and gratitude (as done at YWC); and (4) PLEASE will be supplemented with HALT from 12-step facilitation and additional independent **healthy living modules** exercise, nutrition, and sleep.
- Include distress tolerance DBT skills sessions (STOP, Pros and Cons, ACCEPTS, Self Soothe, TIPP).

We based our groups predominantly on the DBT Skills manual for adolescents, adapted where needed for emerging adults (49).

**Levelling up your health**

Increasing physical activity, improving sleep, and optimizing nutrition are frequently cited in best practice guidelines for various substance use and other psychiatric disorders. Further, these are key components of the DBT emotion regulation skill “PLEASE.” Thus, we created 3 separate groups touching on these topics. A fulsome list of references/resources used for these modules is available upon request.

**Developed, though currently inactive**

Several groups were developed in response to literature reviews and stakeholder consultants, though not implemented to date due to resource constraints regarding clinician time, relatively low group attendance, and prioritizing core evidence-based components (e.g., MET, CBT, MBSR, DBT skills, substance-related health promotion). Groups developed, though not yet implemented, include: 1) **peer space** and 2) **navigating the week**.

**Peer Space** planned to include a drop-in information session and a young adult SMART Recovery group. Mutual support groups for substance use problems have demonstrated effectiveness among adults (50, 51) with promise for adolescents and young adults (20, 45, 52). We created an “Introduction to Mutual Support” group to introduce emerging adults to what mutual support groups are, the different types and their values and structure, and information on how to access/join. We planned to discuss: Alcoholics Anonymous and related “anonymous” groups (e.g., Marijuana Anonymous, Cocaine Anonymous, Narcotics Anonymous, etc.), Loved Ones groups (e.g., Al-Anon, Alateen, and Nar-Anon), Buddhist inspired recovery groups (e.g., Refuse Recovery and Recovery Dharma), Female-identified recovery groups (e.g., Women for Sobriety), and SMART Recovery. We also thought about creating a young-adult specific SMART Recovery group, though need was low and given the broader Adult Concurrent Disorders program and local universities offer weekly SMART Recovery, this was put on hold.

**Navigating the week** was developed to provide a weekly drop-in opportunity to predict and plan for immediate high-risk situations (using relapse prevention techniques) while also promoting and planning substance-free pleasurable activities and substance-related health promotion (like sleep, nutrition, and exercise). This session combined key points from the final core CBT session focused on relapse prevention, key points from leveling up your health, and key points from the drop-in CBT session on planning and scheduling pleasurable activities (which is also discussed during individual MET sessions). Thus, all the components of this group are still incorporated elsewhere in the program, just packaged differently and separately.

**E. Details overview of Loved Ones Education Group**

| **Loved Ones Education Group (CRAFT; implemented)** | |
| --- | --- |
| Session | Mechanisms |
| 1: Group Overview, Safety, and Self Care   - Introducing the group - Discussing problems with loved one and personal wellbeing - Discussing the importance of self-care and promoting self-care as a priority | Communicate empathy, foster a sense of community, and promote self-care |
| 2: Understanding Substance Use   - Psychoeducation about SUDs: definitions, stigma, epidemiology, biopsychosocial risk factors and treatment approaches | Increase insight and perceived self-efficacy |
| 3. Understanding co-occurring mental health concerns   - Psychoeducation about co-occurring mental health concerns: definitions mental health and illness, epidemiology, reasons for comorbidity, biopsychosocial risk factors and treatment approaches, introduction to non-judgmental listening | Increase insight and perceived self-efficacy, improve listening skills |
| 4: Positive Communication   - Discussing communication problems and conversation traps - Positive communication strategies | Improve positive communication and reduce conflict with their loved one |
| 5: Past Patterns & New Strategies   - Understanding the patterns and function of their loved one’s substance use and healthy behaviours (triggers 🡪 behaviour 🡪 consequences) - Introduction to Behaviour makes sense: behavioural formula | Core CBT: Functional Analysis |
| 6: Rewards & Coping with Intoxication   - Behaviour makes sense: behavioural formula - Identifying and planning the delivery of positive attention and other reinforcers - Coping with and responding to substance use behaviour | CBT & Behavioural Economics: Reinforcement to increase behaviours |
| 7: Allowing Negative Consequences   - Allowing and anticipating negative consequences (allowing natural consequences, anticipating negative repercussions) - Behaviour makes sense: behavioural formula | CBT & Behavioural Economics: Consequences to decrease behaviours |
| 8: Talking About Treatment, Review, & Next Steps   - Motivating your loved one to enter treatment (including how to offer advice and collaboratively problem solve) - Session by session review | Improve positive communication  Increase motivation  Mastery and memory consolidation |

**F. Example Clinician Summary**

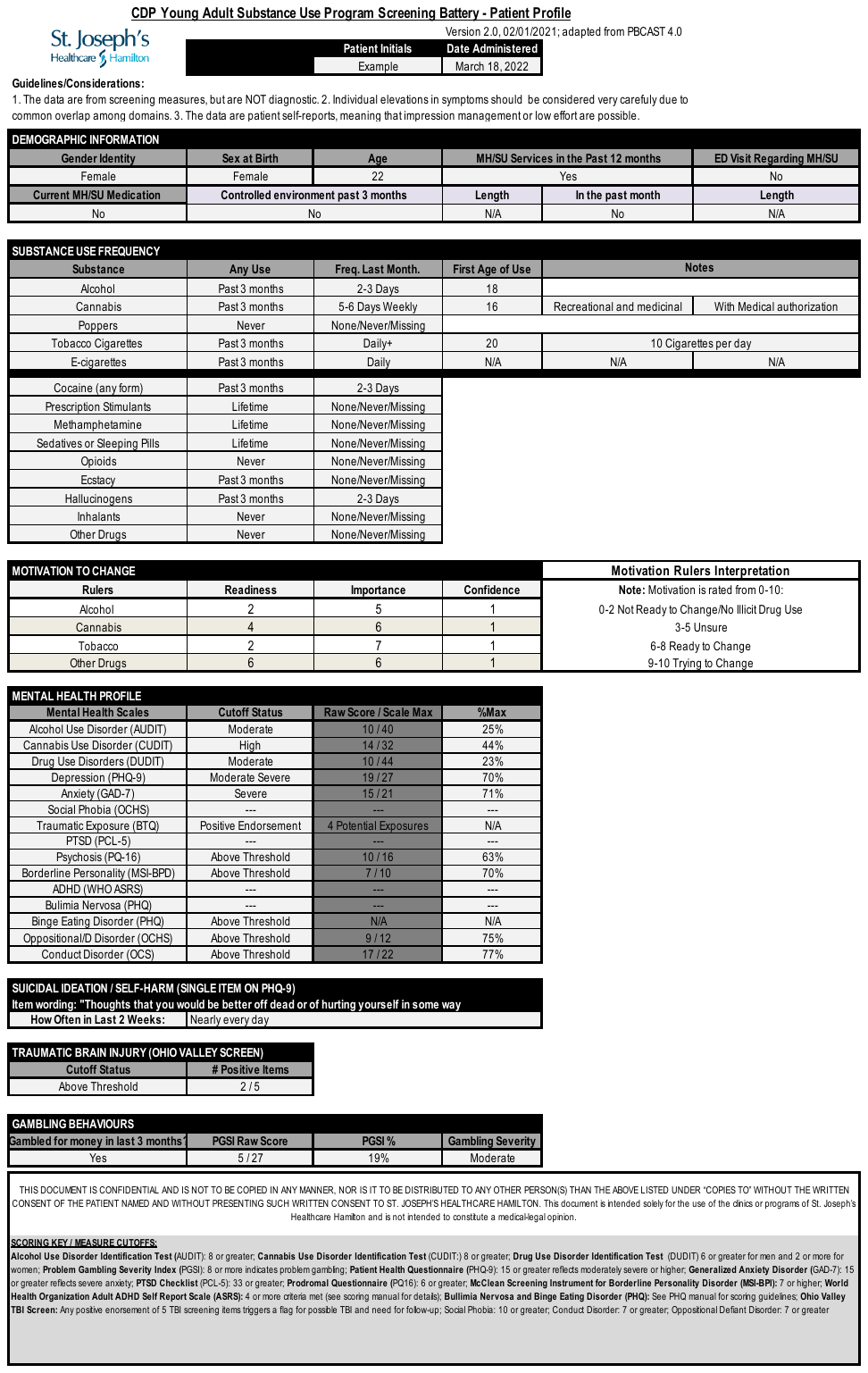

**G. Example MET 1 Personalized Feedback Report**

’

H. Additional Youth Adult Characteristics

Figure H1. Primary and Secondary Reasons for Treatment as Endorsed by Young Adults (n=96).

Primary and Secondary Reasons for Treatment

|  | Primary | Secondary |
| --- | --- | --- |
| Alcohol | 43 | 21 |
| Cannabis | 40 | 18 |
| Cigarettes | 9 | 26 |
| Cocaine | 18 | 8 |
| E-cigarettes | 6 | 16 |
| Sedatives | 5 | 5 |
| Hallucinogens | 2 | 6 |
| Opioids | 3 | 5 |
| Ecstasy | 3 | 2 |
| Stimulants | 1 | 4 |
| Methamphetamine | 3 | 1 |

Figure H2. Average Quality of Life Item Scores at Intake. Means and SDs are presented across items (n=96).

Average Quality of Life Scores

|  |  |
| --- | --- |
| General well-being (emotional, physical, spiritual | 3.77 |
| Day-to-day activities (study, work, leisure, self-care | 3.82 |
| Relationships with friends | 4.52 |
| Relationships with family | 4.38 |
| Coping without substances (deling with life, using your strengths) | 3.63 |

Note: The first 35 participants received the original wording for the coping item that did not specify “without substances.” Clarification was added due to feedback from clinicians. The mean(SD) was 3.9(2.9) before and 3.5(3.0) after the adjustment.

I. AUDIT items

Audit3 to Audit8 response options used in the YA-SUP:

0 – Never

1 – Monthly or less

2 – 2-4 times a month

3 – 2-3 times a week

4 – Daily or almost daily

**Supplementary Material References**

1. Ross E, Vingilis E, Osuch E. An engagement and access model for healthcare delivery to adolescents with mood and anxiety concerns. Early Interv Psychia [Internet]. 2012 Feb [cited 2022 Jul 19];6(1):97-105. Available from: <https://pubmed.ncbi.nlm.nih.gov/22171651/> DOI: <https://doi.org/10.1111/j.1751-7893.2011.00312.x>
2. Dugosh KL, Cacciola JS. Clinical assessment of substance use disorders. In: Saxon AJ, Friedman M, editors. UpToDate. [Internet]. Waltham, MA: UpToDate Inc.; 2019 [updated 2022 Apr 19; cited 2022 Jul 19]. Available from: <https://www.uptodate.com/contents/clinical-assessment-of-substance-use-disorders>.
3. Skewes MC, Gonzalez VM. Principles of addiction: comprehensive addiction behaviours and disorders. Massachusetts: Academic Press; 2013; p. 61-70.
4. Headspace National Youth Mental Health Foundation. Australia: [publisher not found]; c2022. Formulation; [date not found] [cited 2022 Jul 20]. Available from: <https://headspace.org.au/clinical-toolkit/formulation/>
5. Parker A, Hetrick S, Purcell R. Psychosocial assessment of young people – refining and evaluating a youth friendly assessment interview. Aust Fam Physician [Internet]. 2010 Aug [cited 2022 Jul 20];39(8):585. Available from: <https://pubmed.ncbi.nlm.nih.gov/20877754/>
6. Steele DW, Becker SJ, Danko KJ, Balk EM, Adam GP, Saldanha IJ, et al. Brief behavioral interventions for substance use in adolescents: a meta-analysis. Pediatrics [Internet]. 2020 Oct [cited 2022 Jul 20];146(4)e20200351. Available from: <https://pubmed.ncbi.nlm.nih.gov/32928988/> DOI: 10.1542/peds.2020-0351
7. O’Connor EA, Perdue LA, Senger CA, Rushkin M, Patnode CD, Bean SI, et al. Screening and behavioral counseling interventions to reduce unhealthy alcohol use in adolescents and adults: updated evidence report and systematic review for the US Preventive Services Task Force. JAMA [Internet]. 2018 Nov 13 [cited 2022 Jul 20];320(18):1910-28. Available from: <https://jamanetwork.com/journals/jama/fullarticle/2714536> DOI: 10.1001/jama.2018.12086
8. Tanner‐Smith EE, Parr NJ, Schweer‐Collins M, Saitz R. Effects of brief substance use interventions delivered in general medical settings: a systematic review and meta‐analysis. Addiction [Internet]. 2022 Apr [cited 2022 Jul 20]. Available from: <https://pubmed.ncbi.nlm.nih.gov/34647649/> DOI: 10.1111/add.15674
9. Halladay J, Scherer J, Mackillop J, Woock R, Petker T, Linton V, et al. Brief interventions for cannabis use in emerging adults: a systematic review, meta-analysis, and evidence map. Drug Alcohol Depend [Internet]. 2019 Nov 1 [cited 2022 Jul 20];204:107565. Available from: <https://pubmed.ncbi.nlm.nih.gov/31751868/> DOI: 10.1016/j.drugalcdep.2019.107565
10. Meshesha LZ, Soltis KE, Wise EA, Rohsenow DJ, Witkiewitz K, Murphy JG. Pilot trial investigating a brief behavioral economic intervention as an adjunctive treatment for alcohol use disorder. J Subst Abuse Treat [Internet]. 2020 Jun [cited 2022 Jul 20];113:108002. Available from: <https://pubmed.ncbi.nlm.nih.gov/32359674/> DOI: 10.1016/j.jsat.2020.108002
11. Murphy JG, Dennhardt AA, Martens MP, Borsari B, Witkiewitz K, Meshesha LZ. A randomized clinical trial evaluating the efficacy of a brief alcohol intervention supplemented with a substance-free activity session or relaxation training. J Consult Clin Psychol [Internet]. 2019 Jul [cited 2022 Jul 20];87(7):657-69. Available from: <https://pubmed.ncbi.nlm.nih.gov/31070386/> DOI: 10.1037/ccp0000412
12. Carver J, Cappelli M, Davidson S. Taking the next step forward: building a responsive mental health and addictions system for emerging adults [Internet]. Ottawa: Mental Health Commission of Canada; 2015 Jul 17 [cited 2022 May 24]. 108 p. Available from: <https://www.mentalhealthcommission.ca/wp-content/uploads/drupal/Taking%252520the%252520Next%252520Step%252520Forward_0.pdf>
13. Bergman BG, Kelly JF, Nargiso JE, McKowen JW. "The age of feeling in-between": addressing challenges in the treatment of emerging adults with substance use disorders. Cogn Behav Pract [Internet]. 2016 [cited 2022 May 24];23(3):270-88. Available from: <https://psycnet.apa.org/record/2016-02611-001> DOI: 10.1016/j.cbpra.2015.09.008.
14. Ingersoll K. Motivational interviewing for substance use disorders. In: Stein MB, Friedman M, editors. [Internet]. Waltham, MA: UpToDate Inc.; c2020 [updated 2022 Mar 1; cited 2022 Jul 20]. Available from: <https://www.uptodate.com/contents/motivational-interviewing-for-substance-use-disorders>
15. Halladay J, Fein A, MacKillop J, Munn C. PAUSE: the development and implementation of a novel brief intervention program targeting cannabis and alcohol use among university students. Can J Addict [Internet]. 2018 Jun [cited 2022 Jul 20];9(2):34-42. Available from: <https://journals.lww.com/cja/Abstract/2018/06000/PAUSE__The_Development_and_Implementation_of_a.7.aspx> DOI: 10.1097/CXA.0000000000000020
16. Godley SH. The adolescent community reinforcement approach for adolescent cannabis users. 4th ed. Rockville, MD: U.S. Department of Health and Human Services, Substance Abuse and Mental Health Services Administration, Centre for Substance Abuse Treatment; 2001.
17. Kwan B, Rickwood DJ, Telford NR. Development and validation of MyLifeTracker: a routine outcome measure for youth mental health. Psychol Res Behav Manag [Internet]. 2018 Apr 3 [cited 2022 may 26];2018(11):67-77. Available from: <https://www.ncbi.nlm.nih.gov/pmc/articles/PMC5892955/> DOI: 10.2147%2FPRBM.S152342.
18. Burkstein O. Approach to treating substance use disorder in adolescents. In: Brent D, Saxon AJ, Friedman M, editors. [Internet]. Waltham, MA: UpToDate Inc.; 2022 [updated 2022 Jan 21; cited 2022 May 25]. Available from: <https://www.uptodate.com/contents/approach-to-treating-substance-use-disorder-in-adolescents>
19. McKay JR. Psychotherapies for substance use disorders. In: Saxon AJ, Friedman M, editors. [Internet]. Waltham, MA: UpToDate Inc.; 2020 [updated 2020 Sep 28; cited 2022 May 25]. Available from: <https://www.uptodate.com/contents/psychotherapies-for-substance-use-disorders>
20. Kelly JF, Yeterian JD, Cristello JV, Kaminer Y, Kahler CW, Timko C. Developing and testing twelve-step facilitation for adolescents with substance use disorder: manual development and preliminary outcomes. Subst Abuse [Internet]. 2016 Jun 13 [cited 2022 Jul];10:55-64. Available from: <https://www.ncbi.nlm.nih.gov/pmc/articles/PMC4941867/> DOI: 10.4137%2FSART.S39635
21. Webb C. The motivational enhancement therapy and cognitive behavioral therapy supplement: 7 sessions of cognitive behavioral therapy for adolescent cannabis users. Rockville, MD: U.S. Department of Health and Human Services, Substance Abuse and Mental Health Services Administration, Centre for Substance Abuse Treatment; 2002.
22. MacKillop J, Sweet LH, Amlung M. Using neuroeconomics to understand alcohol overvaluation in alcohol use disorder (NEURO ALC) treatment Manual 2019. Unpublished.
23. Drake RE, O'Neal EL, Wallach MA. A systematic review of psychosocial research on psychosocial interventions for people with co-occurring severe mental and substance use disorders. J Subst Abuse Treat [Internet]. 2008 Jan [cited 2022 Jul 21];34(1):123-38. Available from: <https://pubmed.ncbi.nlm.nih.gov/17574803/> DOI: 10.1016/j.jsat.2007.01.011
24. Coco GL, Melchiori F, Oieni V, Infurna MR, Strauss B, Schwartze D, et al. Group treatment for substance use disorder in adults: a systematic review and meta-analysis of randomized-controlled trials. J Subst Abuse Treat [Internet]. 2019 Apr [cited 2022 Jul 21];99:104-16. Available from: <https://pubmed.ncbi.nlm.nih.gov/30797382/> DOI: 10.1016/j.jsat.2019.01.016
25. Silvers JA, Squeglia LM, Thomsen KR, Hudson KA, Ewing SWF. Hunting for what works: adolescents in addiction treatment. Alcohol Clin Exp Res [Internet]. 2019 Apr 1 [cited 2022 Jul 21];43(4):578-92. Available from: <https://www.ncbi.nlm.nih.gov/pmc/articles/PMC6443447/> DOI: 10.1111/acer.13984
26. Casey BJ, Jones RM. Neurobiology of the adolescent brain and behavior: implications for substance use disorders. J Am Acad Child Adolesc Psychiatry [Internet]. 2010 Dec [cited 2022 Jul 21];49(12):1189-201. Available from: <https://pubmed.ncbi.nlm.nih.gov/21093769/> DOI: 10.1016/j.jaac.2010.08.017
27. Pan L, Brent DA. BMJ talk medicine: depression in children [podcast on the Internet]. London: BMJ Best Practice; 2020 [cited 2022 Jul 21]. Available from: <https://soundcloud.com/bmjpodcasts/depression-in-children>
28. Creswell C, Waite P, Cooper PJ. Assessment and management of anxiety disorders in children and adolescents. Arch Dis Child [Internet]. 2014 Jul [cited 2022 Jul 21];99(7):674-8. Available from: <https://pubmed.ncbi.nlm.nih.gov/24636957/> DOI: 10.1136/archdischild-2013-303768
29. Rush AJ. Unipolar major depression in adults: choosing initial treatment. In: Roy-Byrne PP, Solomon D, editors. UpToDate. [Internet]. Waltham, MA: UpToDate Inc.; 2020. [updated 2020 Nov 18; cited 2022 Jul 21]. Available from: <https://www.uptodate.com/contents/unipolar-major-depression-in-adults-choosing-initial-treatment>
30. MacKinnon DF. BMJ talk medicine: depression in adults [podcast on the Internet]. London: BMJ Best Practice; 2021 [cited 2022 Jul 21]. Available from: <https://soundcloud.com/bmjpodcasts/depression-in-adults>
31. Gorelick DA. Cannabis use disorder in adults. In: Saxon AJ, Friedman M, editors. UpToDate. [Internet]. Waltham, MA: UpToDate Inc.; 2021 [updated 2021 Dec 16; cited 2022 Jul 21]. Available from: <https://www.uptodate.com/contents/cannabis-use-disorder-in-adults>
32. Sampl S, Kadden R. Motivational enhancement therapy and cognitive behavioral therapy for adolescent cannabis users: 5 sessions. Rockville, MD: U.S. Department of Health and Human Services, Substance Abuse and Mental Health Services Administration, Centre for Substance Abuse Treatment; 2001.
33. Costa el Hage M, Wolfe J, Henderson J. C-SMART Revised Manual - 2007. Unpublished.
34. Kadden R, Carroll K, Donovan D, Cooney N, Monti PM, Abrams D, et al. Cognitive-behavioral coping skills therapy manual: a clinical research guide for therapists treating individuals with alcohol abuse and dependence. 3rd ed. Rockville, MD: US Department of Health and Human Services, Substance Abuse and Mental Health Services Administration, Centre for Substance Abuse Treatment; 1995.
35. Monti PM, Rohsenow DJ. Coping-skills training and cue-exposure therapy in the treatment of alcoholism. Alcohol Res Health [Internet]. 1999 Nov 2 [cited 2022 Jul 21];23(2):107-15. Available from: <https://pubmed.ncbi.nlm.nih.gov/10890804/>
36. Herie M, Watkin-Merek L. Structured relapse prevention concurrent disorders: an outpatient counselling approach. 2nd ed. Hamilton, ON: Centre for Addiction and Mental Health; 2006.
37. Milosevic I, Chudzik S, Boyd S, McCabe, R, Aiken A, Nault J. Crossing paths: cognitive behavioural therapy for depression, anxiety & substance use problems – group manual. 2019. Unpublished.
38. Miller WR, editor. Combined behavioral intervention manual: a clinical research guide for therapists treating people with alcohol abuse and dependence. Rockville, MD: US Department of Health and Human Services; 2004.
39. Hatchard T, Hames J, Ladak A, Laman J. Mind vs mood: changing our thoughts & behaviours using CBT strategies – client manual. 2020. Unpublished.
40. Litt MD, Kabela-Cormier E, Kadden RM. Network support for alcohol treatment: mechanisms and effectiveness. c2009. Unpublished.
41. Korecki JR, Schwebel FJ, Votaw VR, Witkiewitz K. Mindfulness-based programs for substance use disorders: a systematic review of manualized treatments. Subst Abuse Treat Prev Policy [Internet]. 2020 Jul 29 [cited 2022 Jul 22];15(1):51. Available from: <https://pubmed.ncbi.nlm.nih.gov/32727559/> DOI: 10.1186/s13011-020-00293-3
42. Sancho M, De Gracia M, Rodriguez RC, Mallorquí-Bagué N, Sánchez-González J, Trujols J, et al. Mindfulness-based interventions for the treatment of substance and behavioral addictions: a systematic review. Front Psychiatry [Internet]. 2018 Mar 29 [cited 2022 Jul 22];9:95. Available from: <https://pubmed.ncbi.nlm.nih.gov/29651257/> DOI: 10.3389/fpsyt.2018.00095
43. Breedvelt JJF, Amanvermez Y, Harrer M, Karyotaki E, Gilbody S, Bockting CL, et al. The effects of meditation, yoga, and mindfulness on depression, anxiety, and stress in tertiary education students: a meta-analysis. Front Psychiatry [Internet]. 2019 Apr 24 [cited 2022 Jul 22];10:193. Available from: <https://pubmed.ncbi.nlm.nih.gov/31068842/> DOI: 10.3389/fpsyt.2019.00193
44. Halladay JE, Dawdy JL, McNamara IF, Chen AJ, Vitoroulis I, McInnes N, et al. Mindfulness for the mental health and well-being of post-secondary students: a systematic review and meta-analysis. Mindfulness. 2018 Jun 28 [cited 2022 Jul 22];10(3):397-414. Available from: <https://www.semanticscholar.org/paper/Mindfulness-for-the-Mental-Health-and-Well-Being-of-Halladay-Dawdy/0e0072838f8806a60d3e1b3c47074a2ac28407a8> DOI: 10.1007/S12671-018-0979-Z
45. Fadus MC, Squeglia LM, Valadez EA, Tomko RL, Bryant BE, Gray KM. Adolescent substance use disorder treatment: an update on evidence-based strategies. Curr Psychiat Rep [Internet]. 2019 Sep 14 [cited 2022 Jul 22];21(10):96. Available from: <https://www.ncbi.nlm.nih.gov/pmc/articles/PMC7241222/> DOI: 10.1007%2Fs11920-019-1086-0
46. Broderick PC. Learning to breathe: a mindfulness curriculum for adolescents to cultivate emotion regulation, attention, and performance. 2nd ed. Oakland, CA: New Harbinger Publications; 2021.
47. Giannelli E, Gold C, Bieleninik L, Ghetti CM, Gelo OCG. Dialectical behaviour therapy and 12‐step programmes for substance use disorder: a systematic review and meta‐analysis. Couns Psychother Res [Internet]. 2019 May [cited 2022 Jul 22];19(3):274-85. Available from: <https://www.researchgate.net/publication/333480402_Dialectical_behaviour_therapy_and_12-step_programmes_for_substance_use_disorder_A_systematic_review_and_meta-analysis> DOI: 10.1002/capr.12228.
48. Cavicchioli M, Movalli M, Vassena G, Ramella P, Prudenziati F, Maffei C. The therapeutic role of emotion regulation and coping strategies during a stand-alone DBT Skills training program for alcohol use disorder and concurrent substance use disorders. Addict Behav [Internet]. 2019 Nov [cited 2022 Jul 22];98:106035. Available from: <https://pubmed.ncbi.nlm.nih.gov/31302312/> DOI: 10.1016/j.addbeh.2019.106035
49. Rathus JH, Miller AL. DBT skills manual for adolescents. New York: Guilford Publications; 2014.
50. Beck AK, Forbes E, Baker AL, Kelly PJ, Deane FP, Shakeshaft A, et al. Systematic review of SMART recovery: outcomes, process variables, and implications for research. Psychol Addict Behav [Internet]. 2017 Feb [cited 2022 Jul 22];31(1):1-20. Available from: <https://pubmed.ncbi.nlm.nih.gov/28165272/> DOI: 10.1037/adb0000237
51. Kelly JF, Abry A, Ferri M, Humphreys K. Alcoholics anonymous and 12-step facilitation treatments for alcohol use disorder: a distillation of a 2020 Cochrane review for clinicians and policy makers. Alcohol Alcoholism [Internet]. 2020 Jul 6 [cited 2022 Jul 22];55(6):641-51. Available from: <https://academic.oup.com/alcalc/article/55/6/641/5867689> DOI: 10.1093/alcalc/agaa050
52. Bergman BG, Kelly JF, Fallah-Sohy N, Makhani S. Emerging adults, mutual-help organizations, and addiction recovery: what does the science tell us? In: Smith DC, editor. Emerging adults and substance use disorder treatment: developmental considerations and innovative approaches. Oxford: Oxford University Press; 2018. p. 167-95.
